# A novel approach for evaluating contact patterns and risk mitigation strategies for COVID-19 in English Primary Schools with application of Structured Expert Judgement

**DOI:** 10.1101/2020.08.13.20170068

**Authors:** R.S.J. Sparks, W.P. Aspinall, E. Brooks-Pollock, R.M. Cooke, L. Danon, J. Barclay, J.H. Scarrow, J.H. Cox

**Affiliations:** School of Earth Sciences, University of Bristol, Bristol BS8 1RJ, United Kingdom; Aspinall and Associates, Tisbury SP3 6HF, United Kingdom; School of Veterinary Sciences, University of Bristol, Office OF24, Churchill Building, Langford, Bristol BS40 5DU, United Kingdom; Institute of Applied Mathematics, Delft University of Technology, Building 28, Mourik Broekmanweg 6, 2628 XE Delft, Netherlands; Department of Computer Sciences, College of Engineering, Mathematics and Physical Sciences, Harrison Building, Streatham Campus, University of Exeter, North Park Road, Exeter EX4 4QF, United Kingdom; School of Environmental Sciences, University of East Anglia, Norwich Research Park, Norwich NR4 7TJ, United Kingdom; Departamento de Mineralogía y Petrologia, Facultad de Ciencias, Universidad de Granada, 18071 Granada, Spain; The Royal Society, 6-9 Carlton House Terrace, London SW7 5QR, United Kingdom

## Abstract

**Background:** Contact patterns are the drivers of close-contacts infections, such as COVID-19. In an effort to control COVID-19 transmission in the UK, schools were closed on 23 March 2020. With social distancing in place, Primary Schools were partially re-opened on 1 June 2020, with plans to fully re-open in September 2020. The impact of social distancing and risk mitigation measures on children’s contact patterns is not known.

**Methods:** We conducted a structured expert elicitation of a sample of Primary Headteachers to quantify contact patterns within schools in pre-COVID-19 times and how these patterns were expected to change upon re-opening. Point estimates with uncertainty were determined by a formal performance-based algorithm. Additionally, we surveyed school Headteachers about risk mitigation strategies and their anticipated effectiveness.

**Results:** Expert elicitation provides estimates of contact patterns that are consistent with contact surveys. We report mean number of contacts per day for four cohorts within schools along with a range at 90% confidence for the variations of contacts among individuals. Prior to lockdown, we estimate that, mean numbers per day, younger children (Reception and Year 1) made 15 contacts [range 8..35] within school, older children (Year 6) 18 contacts [range 5.. 55], teaching staff 25 contacts [range 4.. 55) and non-classroom staff 11 contacts [range 2.. 27]. Compared to pre-COVID times, after schools re-opened the mean number of contacts were reduced by about 53% for young children, about 62% for older children, about 60% for classroom staff and about 64% for other staff. Contacts between teaching and non-teaching staff reduced by 80%, which is consistent with other independent estimates. The distributions of contacts per person are asymmetric indicating a heavy tail of individuals with high contact numbers.

**Conclusions:** We interpret the reduction in children’s contacts as a consequence of efforts to reduce mixing with interventions such as forming groups of children (bubbles) who are organized to learn together to limit contacts. Distributions of contacts for children and adults can be used to inform COVID-19 transmission modelling. Our findings suggest that while official DfE guidelines form the basis for risk mitigation in schools, individual schools have adopted their own bespoke strategies, often going beyond the guidelines.

## Introduction

On 11^th^ May 2020 Prime Minister Boris Johnson announced that selected primary age children would return to school on 1^st^ June in England. The returning cohort would include Reception, Year 1 and Year 6, noting that there would also be Year 2 to 5 children of frontline workers and those identified as vulnerable were already being taught in many schools. Nursery age children were also invited to return with some nurseries being part of Primary schools. The devolved administrations decided at the time not to re-open schools. The Government abandoned the proposal for a full return of Primary school children in England before the summer holidays. There is now an expectation that schools will fully re-open in September.

The partial re-opening of Primary schools has been widely debated with concerns from some parents, teaching unions and teacher associations about the safety of the children and school staff. There was also concern about the effect on infection rate in the wider community, for example by triggering a second wave. Some schools re-opened on 1^st^ June, some have delayed their re-start while, in other cases, schools have not reopened under advice of local education authorities. Data from DfE indicates on 15^th^ July that about 88% of state-funded Primary Schools have re-opened to some extent. The community response has been variable and between 1^st^ and 15^th^ June 2020 approximately one third of eligible children returned. The numbers have increased somewhat and were 41% (year 1) and 49% (year 6) on 2^nd^ July. Between 18^th^ May and 31^st^ July 2020 there have been 247 COVID-19 related incidents in schools of which 116 were tested to be positive test (PHE 2020).

Contact patterns, as drivers of transmission for close-contact infections, are an essential component of epidemiological models (Del Valle et al. 2007; Mossong et al. 2008; Eames et al. 2012; Danon et al. 2013; Zhang et al. 2020). There is, however, a paucity of information about the contact patterns of younger children, especially in school settings, due to the challenges involved in collecting data from children. Here we address this question by applying Structured Expert Judgement (SEJ) using leaders in Primary Schools as experts. SEJ is a well-established approach to quantifying parameters and their attendant uncertainties where there are no data or the data are sparse or of poor quality or are highly empirical in character or have large associated uncertainties (Cooke 1991; Colson & Cooke 2017). This is the case for many epidemiological parameters, including contact patterns which are used in combination with the probability of transmission and the infectious period to describe the reproduction number in a population. SEJ has been widely applied to risk assessment and uncertainty analysis in many areas of science, engineering, the environment, business and public health (Colson & Cooke 2017). The Classical Model for SEJ has been deployed in several public health policy applications (e.g. Barons & Aspinall 2020; Beshearse et al. in press; Hald et al. 2016; Hincks et al. 2006; Martell et al. 2007; Tyshenko et al. 2012, 2016). Notably SEJ is not widely used in epidemiology.

We identified 34 volunteers from within the Royal Society Schools network, most of whom had a STEM background. The group of experts were leaders and senior staff of state-funded Primary Schools in England. The focus of the SEJ was to quantify contact patterns within schools and investigate how these patterns had changed between pre-COVID and COVID times. We also took the opportunity to survey school staff about their risk mitigation strategies and asked for information to assess their effectiveness. Under current circumstances the usual approach to SEJ involving typically 1 or 2 days of face-to-face discussions between experts was impractical and would in any case contravene social distancing requirements. We therefore augmented the SEJ and survey with six structured interviews to improve our understanding of the responses.

## Methods

To determine variable or parameter uncertainty distributions we utilized a validated elicitation method, Classical Model for Structured Expert Judgement (SEJ). The method is described in Cooke (1991), Colson & Cooke (2017) and in: https://en.wikipedia.org/wiki/Structured_expert_judgment:_the_classical_model. The distinctive feature of the Classical Model is that experts, in a group being elicited, are empirically calibrated for their ability to judge uncertainties in terms of statistical accuracy taken over a set of knowable test variables from within the subject matter domain. The resulting performance-based calibration scores are used as weights to create a synthetic combined distribution which is also scored for statistical accuracy and informativeness. The so-called Decision Maker (DM) is in effect, a synthesized pseudo-expert, which represents the group’s collective judgement. Expert weights are predicated on the mathematical concept of ‘proper scoring’ rules, with the resulting Decision Maker solutions providing a rational consensus of the experts’ judgments, weighted according to the individual calibration performance scores. From here in the article the volunteer primary school staff will be termed experts.

Calibration questions, to which true answers are known post hoc, for the SEJ were prepared by WPA. Calibration and target item elicitation questionnaires are provided in Appendix 1. Elicitation questions were devised by the authors to elucidate contact patterns within schools. We also took the opportunity to ask the Head Teachers about risk mitigation measures that they have put in place. Before circulating to the experts the calibration and elicitation questions were reviewed by Professor Andrew Noyes (Faculty of Education, University of Nottingham) to check their clarity. Some revisions were made as a consequence of these reviews. Normally experts are convened at least in one or more face-to-face meetings in an SEJ. A meeting usually covers: introduction to the methodology; calibration of the experts using seed questions; presentation and discussion of the questions; and a time for the experts to answer the questions. A meeting might last 1 or 2 days and with plenty of time for discussion on the evidence that informs responses to the questions. After the data have been processed there then an opportunity to discuss the results and there may be opportunities to re-elicit some questions.

During national lockdown and with experts distributed in Primary Schools across England the normal procedure was not possible. A briefing session was held by zoom and most of the experts participated. The briefing session was recorded enabling those who could not attend the chance to hear the proceedings. For both the seed and elicitation questions Aspinall and Sparks were available to respond to queries and clarifications by email. To compensate for the lack of a meeting six experts were chosen for structured interviews (Longhurst, 2015). The question protocol ensured participants described the thinking behind their quantitative answers but also allowed a free exploration of topics the headteachers perceived of relevance to adult and child contacts.

## Data Resources

### National Data

We extracted data on primary schools in England from DfE links. Information figures for 2019 and 2018 can be found at:

https://assets.publishing.service.gov.uk/government/uploads/system/uploads/attachment_data/file/826252/Schools_Pupils_and_their_Characteristics_2019_Accompanying_Tables.xlsx

https://www.besa.org.uk/key-uk-education-statistics/

https://www.gov.uk/government/statistics/school-workforce-in-england-november-2018

The basic facts about state-funded primary schools in England are: 16,769 schools; 4,727,090 pupils; 216,500 teachers; 176,679 teaching assistants; and 132,085 other support staff.

Data for daily national attendance of pupils, teachers, teaching assistants and ancillary staff in schools can be found at:

https://www.gov.uk/government/collections/attendance-in-education-and-early-years-settings-during-the-coronavirus-covid-19-outbreak

### The study schools

The major source of data comes from the expert elicitation. The Royal Society manages a Schools network consisting of 900 schools across the four nations of the UK and the network includes 1300 teachers. The focus of the network is STEM education and so the majority of the teachers within the network have a STEM background. A call was made for volunteers to participate as experts to help characterise contact data between children, teachers and other staff in schools. The STEM background of the volunteers was an advantage for eliciting numerical data framed by basic statistical concepts with which the teachers were already familiar. From 36 initial volunteers, although 2 dropped out due to being in Wales or Scotland. Of the remaining 34, the 1^st^ questionnaire was completed by 33 teachers. The 2^nd^ questionnaire and calibration questions were completed by 26 teachers and calibration questions were completed by 28 teachers.

The Primary Schools ranged in size from 65 pupils to 910 with an average of 376 children. The national average is 282 pupils. The schools are geographically well distributed: Eastern England (4); South West (8); London (5); North West (5); Midlands (5); South and South East (6); North East (1). The schools are a mixture of urban and rural settings. Teaching staff ranged from 53 to 5 teachers (average 18) and support staff ranged from 66 to 4 (average 24). The national average number of teachers and support staff per primary school are 13 and 18 respectively. These data indicate that the study group of schools is nationally representative. All the recruited experts were in positions of senior management or authority with descriptions including: Head or Deputy Head Teachers (21); Head of Department or subject co-ordinator (8); regional or area Mentor (5).

We divided the people in the schools into four cohorts based on the children’s Year groups and staff roles. While a simplification, the cohorts are expected to have different contact characteristics, e.g. with respect to interactions between children, between children and adults and between adults. The cohorts are as follows: *Cohort 1* are Nursery, Reception and Year 1 children; *Cohort 2* are Years 2-6 children, noting that Year 2-5 children are those of key workers and from vulnerable environments; *Cohort 3* are classroom teachers and teaching assistants; *Cohort 4* are non-teaching staff such as administrators, cooks, etc, some of whom are expected to have more limited contact with children.

The expert elicitation was conducted under a protocol of confidentiality and non-attribution in order to encourage individual participants to express their own professional judgements about contact patterns, and to remove any constraint, such as expressing only official or policy expectations.

## Elicitation Results

### The experts

The elicitation produced 26 complete responses from experts who had undergone Classical Model (CM) calibration process. Thus, each had a personal statistical accuracy score and an information score (Table 1). These scores are the basis for ascribing a relative performance weight to each participant for pooling judgements in the context of enumerating uncertainty assessments for specific target/query items. Classical Model non-optimised ltem weights combination solutions (i.e. all teachers given some weight based on calibration scores) are listed in Table 1^1^. These weights are not the same as ascribing equal weights to all participants. When combining judgements collectively, each person’s uncertainty distribution is weighted according as their own performance score, so each contributes with some real positive weight to the overall outcome.

**Table 1.** Elicitation experts’ calibration

| Expert | Statistical<br>accuracy p-<br>value | Relative<br>information<br>metric | No.<br>seed<br>items | Relative<br>performanc<br>e score | Normalised<br>weight with<br>DM |
| --- | --- | --- | --- | --- | --- |
| E01 | 8.92E-03 | 2.342 | 7 | 1.29E-2 | 1.55E-2 |
| E02 | 1.60E-03 | 2.262 | 7 | 2.83E-3 | 3.39E-3 |
| E04 | 7.07E-04 | 2.597 | 7 | 1.00E-3 | 1.20E-3 |
| E05 | 1.89E-04 | 2.334 | 7 | 2.79E-4 | 3.35E-4 |
| E07 | 1.62E-02 | 2.328 | 7 | 1.95E-2 | 2.34E-2 |
| E08 | 1.24E-02 | 2.317 | 7 | 2.21E-2 | 2.65E-2 |
| E09 | 1.82E-01 | 1.574 | 7 | 1.79E-1 | 2.15E-1 |
| E10 | 1.82E-01 | 2.204 | 7 | 9.21E-2 | 1.10E-1 |
| E11 | 1.24E-02 | 2.331 | 7 | 1.20E-2 | 1.44E-2 |
| E12 | 4.13E-09 | 2.639 | 7 | 1.27E-8 | 1.52E-8 |
| E13 | 2.45E-07 | 2.060 | 7 | 3.78E-7 | 4.53E-7 |
| E17 | 7.07E-04 | 1.622 | 7 | 9.07E-4 | 1.09E-3 |
| E18 | 1.62E-02 | 2.239 | 7 | 2.01E-2 | 2.41E-2 |
| E19 | 7.07E-04 | 2.410 | 7 | 1.36E-3 | 1.63E-3 |
| E20 | 1.24E-02 | 2.220 | 7 | 2.45E-2 | 2.94E-2 |
| E21 | 2.01E-02 | 2.231 | 7 | 2.83E-2 | 3.40E-2 |
| E22 | 7.07E-04 | 1.398 | 7 | 7.92E-4 | 9.50E-4 |
| E23 | 6.52E-05 | 2.050 | 7 | 8.92E-5 | 1.07E-4 |
| E24 | 1.24E-02 | 2.383 | 7 | 1.47E-2 | 1.76E-2 |
| E25 | 1.89E-04 | 1.650 | 7 | 3.06E-4 | 3.67E-4 |
| E26 | 2.01E-02 | 2.133 | 7 | 2.28E-2 | 2.74E-2 |
| E28 | 1.62E-02 | 1.944 | 7 | 2.22E-2 | 2.67E-2 |
| E30 | 1.42E-01 | 2.184 | 7 | 9.95E-2 | 1.19E-1 |
| E32 | 2.01E-02 | 2.634 | 7 | 1.89E-2 | 2.27E-2 |
| E33 | 1.52E-03 | 2.221 | 7 | 1.74E-3 | 2.09E-3 |
| E35 | 1.52E-03 | 2.322 | 7 | 2.55E-3 | 3.06E-3 |
| E36 | 1.89E-04 | 2.176 | 7 | 1.47E-4 | 1.77E-4 |
| **Solution**<br>**DM** | **5.33E-01** | **0.813** |  | **2.33E-1** | **2.79E-1** |

The calibration “*p*-value” score is calculated on the basis of a Chi-squared test on the deviations of the expert’s judgements over the seed questions, relative to the known values used for calibration. Using Shannon’s relative information statistic, the Chi-squared test takes the frequencies with which the expert’s assessed calibration variable values fall within various ranges, and compares these with the counts of actual (known) item values in the same ranges; this produces relative probabilities of match per item, which can be summed over all calibration items to form a measure of the expert’s statistical accuracy. When combined with the expert’s information metric (see Cooke, 1991) in a product, this *p*-value provides the ‘statistical accuracy’ part of the expert’s performance score. In effect, a calibration *p*-value can be thought equivalent to the probability that an expert’s performance would be regarded as statistically accurate by chance but, in the Classical Model, it is not used in the sense of an hypothesis test. Rather, it is the formal mathematical basis for enumerating the metric for the statistical accuracy of the expert’s assessments and is used, with a companion information metric, for scoring performance in assessing uncertainties. A high *p*-value indicates a close correspondence between the expert’s assessment values and the known values (a perfect score would be *p* = 1); a very low *p*-value signals major deviations exist between the assessed and actual values. The calibration metric is a ‘fast’ function and typically changes markedly between experts in an elicitation.

To provide some context for the teachers’ panel revealed performance scores, the calibration and weighting profile of the group is very similar to that of other professional expert elicitations; for example, in Figure 1 a profile is shown for a medical panel with the same number of participants (n.b. some points overlap). While the overall range of the teachers’ statistical accuracy *p*-values - from highest to lowest - is not atypical, in this case there are fewer individuals with very low *p*-values (i.e. *p* < 10^-6^) than in the medical panel; the latter represents the more usual case. With the exception of one teacher (a very low *p*-value outlier not shown here), the majority of the teachers’ *p*-values are more clustered at higher *p*-values than those obtained for the medical panel. On the other hand, the relative information scores of the teachers are generally lower than those of the medical experts; this may reflect inherent differences in the natures of the precision of the data types each group were considering.

Our conclusion is that the teachers’ elicitation does not exhibit any substantive shortcomings compared to other cases, notwithstanding it was conducted with minimal briefing and without the benefit of a plenary workshop to help focus judgements. In short, the Head teachers’ performances proved to be as strong, collectively, as those of many other groups of experts (Colson and Cooke 2017, for other case profiles).

**Figure 1.**
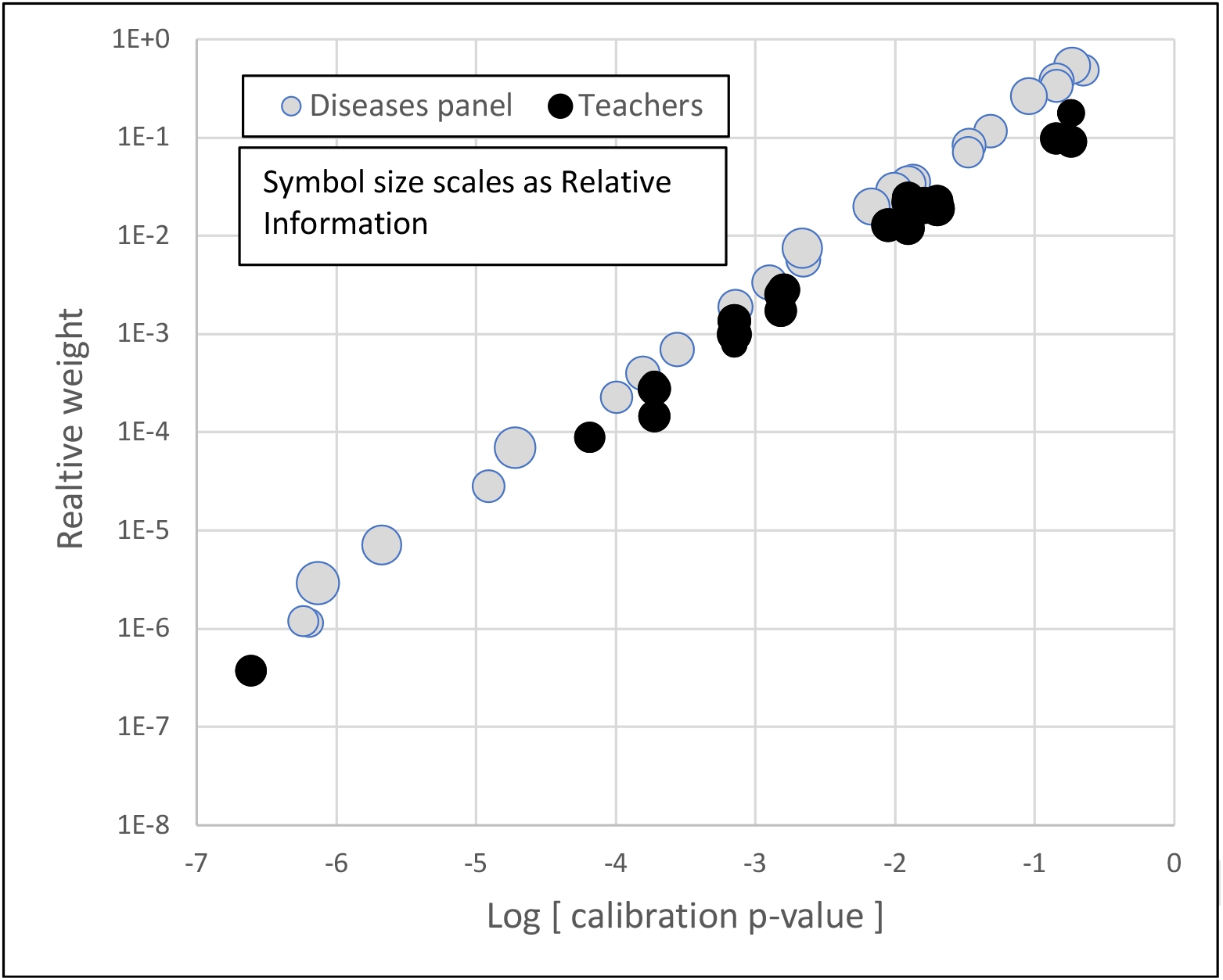
Teachers’ group calibration and weights profile, compared with a childhood diseases medical experts panel of the same size. Each panel comprised 26 participants; the two panels were scored on different subject-matter calibration questions. The plot shows individuals with calibration *p*-value at least 10^-7^, and symbol size scales as the expert’s Relative Information score; there are several over-lapping points.

The combination Decision Maker (DM) -- the combination of all experts -- is statistically more accurate than any individual expert. The range of relative information metric scores is also typical; this is a ‘slow’ function that does not vary greatly from one expert to another. Higher values indicate experts who provided tighter (more informative) uncertainty ranges. However, there is here, and usually, an inverse relationship between informativeness and statistical accuracy: experts who are too narrow or too precise with their uncertainty judgements tend to ‘miss the target’ too frequently and their performance scores are penalised as a consequence. On the other hand, the goal is to identify those experts who are both informative and statistically accurate. This is usually a minority of a group, and such persons are not identifiable *a priori* on common grounds such as professional standing; the best uncertainty assessors are discovered only after calibration. The DM has a low information score compared to individual experts, and this is the price paid for the DM’s superior statistical accuracy. As noted above, it is well-established that individual experts tend to under-estimate true variable uncertainties (Lin & Bier 2008), and pooling via the DM redresses this trait. The combination DM out-performs the best single expert and provides a set of target item solutions that is superior to any single expert.

Individual expert’s relative performance scores (column 5 in Table 1) are computed from the products of their *p*-value and information scores per item, and are unnormalised. When the DM is included as a synthetic expert, performance scores are adjusted and normalised to sum to unity across the group (with DM included). These are the relative weights (column 6 in Table 1) that are used in the Classical Model for combining judgements on target items.

Two experts (E09 & E10) jointly achieve the best statistical accuracy scores (0.182); their judgment influences are differentiated in the analysis by their different information scores - one is more informative than the other and is consequently rewarded with a marginally higher overall weight than the other (see two rightmost black points on Figure 1).

Prior to the main elicitation and before 1^st^ June we asked the experts to forecast the proportion of returning pupils and teachers on two dates (1^st^ and 15^th^ June). When completing these forecasts some teachers indicated that their estimates and ranges were based on surveys of parents before 1 June, conducted for planning purposes. As such, these percentages did not represent personal judgements, *sensu stricto*. However, absent comments from other teachers, it was not possible to know how general pre-return surveys were, or how many respondents had provided percentages data based on similar surveys. Thus, for the purposes and goal of our elicitation exercise, we chose to regard all the inputs on the percentages of pupils returning as representing objective, informed judgements and treated them uniformly when processing the entire group’s responses.

The forecasts were received before or by 27^th^ May 2020. There was a wide range of responses (Figure 2 and Table 2), indicating a remarkable diverse set of circumstances in individual schools and community enthusiasm or lack of enthusiasm for a return to school. This was borne out by the expert interviews where the actual return had been highly variable across just six schools driven both by community perception and school’s mitigation measures. However, when averaged over all of the teachers, the median forecasts are very close (indeed, for pupil’s attendance, identical) to the national attendance on 1^st^ June (Table 2). The national attendance on 15^th^ June was similar to 1^st^ June but had increased by 2^nd^ July to levels similar to those anticipated by the teachers for 15^th^ June. These results demonstrate the ability of the teachers, as experts, to make good forecasts and strengthen the belief that the study schools are a dependable representative sample of primary schools in England.

**Table 2.**
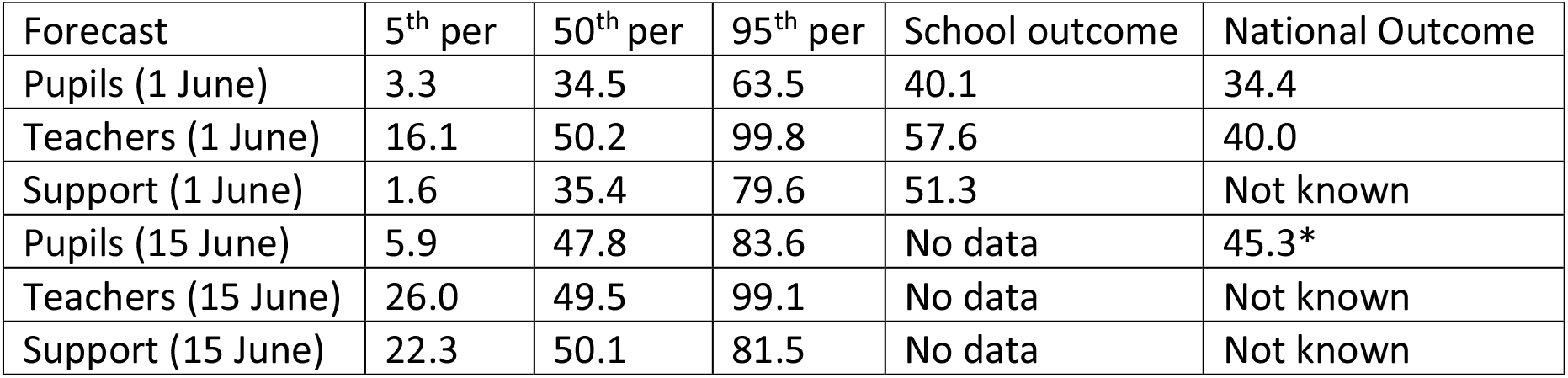
Forecasts as percentages of return to school by experts. Date for pupils are for total reception, Year 1 and Year 6 children. * data refer to 2^nd^ July.

**Figure 2.**
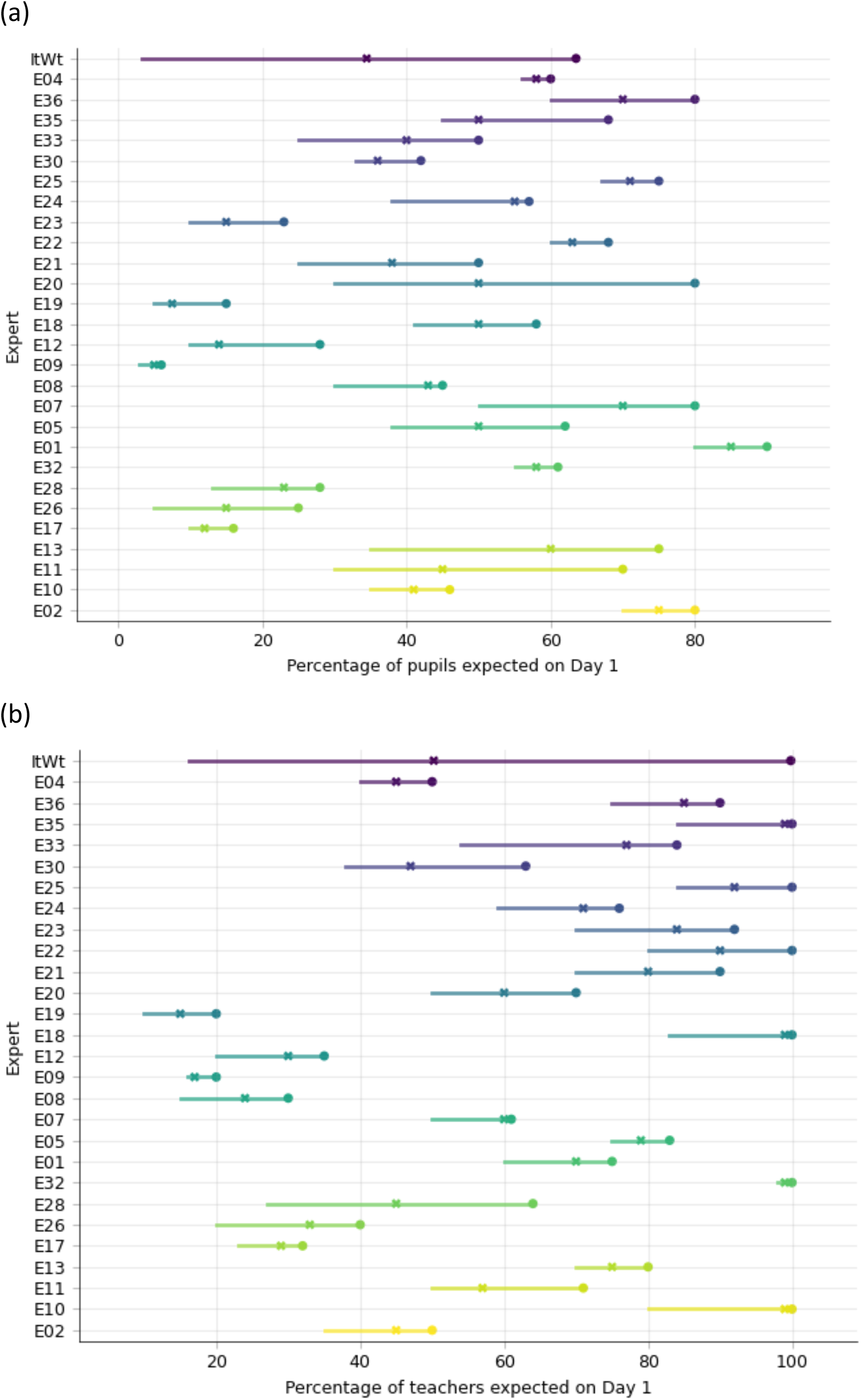
Forecasts of numbers of pupils (a) and teachers (b) returning to school on 1^st^ June 2020. Forecast returns were received before or by 24 May 2020.

The six structured interviews strengthened the assessment of the teachers as thoughtful and careful in thinking through the responses to the questions even thought they were regarded as challenging. All thought about variations between different children, differences in the school day, differences between class time and breaks, and in roles and interactions of staff. Two of the interviewees had consulted with other staff to help them think about contacts.

### Elicitation data

Here we present and interpret the quantitative responses from the elicitation. Table 3 itemises the questions for which quantitative answers were requested. The questions largely focus on contacts between persons in schools as epidemiological models use contact data as a basis for modelling transmission of infection (Danon et al. 2013). Essentially the greater the number of contacts and the longer the duration of those contacts the greater the chance of infection transmission.

**Table 3.** Quantitative elicitation questions

| Number | Question |
| --- | --- |
| 1b | *If you use bubbles please describe the number of pupils in a bubble and the approximate spacing between bubbles during class time. The opportunity is given to give separate answers for Cohort 1 and Cohort 2.* |
| 2a | *How many people does a typical<sup>11</sup> Cohort 1 child come into face to face contact with (conversation within 1 metre for 5 minutes or more) on a <u>normal</u> school day in a covid free world?* |
| 2b | *Thinking about the behaviour of individual children<sup>12</sup> give a range of contacts for Cohort 1 children around your central answer to Q2a for <u>normal</u> times.* |
| 3a | *How many people does a typical Cohort 1 child come into face to face contact with (conversation within 1 metre for 5 minutes or more) on a <u>new normal</u> school day?* |
| 3b | *Thinking about the behaviour of individual children give a range of contacts for Cohort 1 children around your central answer to Q3a for <u>new normal</u> times.* |
| 4 | *Do you think that there is any significant difference<sup>13</sup> in the contacts made by nursery, reception and year 1 children? If yes, please indicate the likely difference in contact <u>referenced to reception age children</u>. Put a percentage to indicate a difference (either way); for example you might judge that year 1 children have 80% of the contacts of a reception child.* |
| 5a | *How many people does a typical Cohort 2 child come into face to face contact with (conversation within 1 metre for 5 minutes or more) on a <u>normal</u> school day?* |
| 5b | *Thinking about the behaviour of individual children, give a range of contacts for Cohort 2 children around your central answer to Q5a for <u>normal</u> times.* |
| 6a | *How many people does a typical Cohort 2 child come into face to face contact with (conversation within 1 metre) on a <u>new normal</u> school day?* |
| 6b | *Thinking about the behaviour of individual children, give a range of contacts for Cohort 2 children around your central answer to Q6a for <u>new normal</u> times.* |
| 7 | *Do you think that there is any difference in the contacts made by Year 2-5 children compared to Year 6<sup>14</sup>. If yes please indicate the likely difference in contact referenced to year 6 age children. Put a percentage to indicate a difference (either way); for example you might judge that year 2 to 5 children have 120% or 80% of the contacts of a Year 6 child.* |
| 8a | *How many people (both children and other adults) does a Cohort 3 adult come into face to face contact (within 1 metre for 5 minutes or more) with on a <u>normal</u> school day?* |

<sup>11</sup> In this and subsequent similar questions you are being asked about the typical or average behaviour and your uncertainty in this average or typical behaviour.
<sup>12</sup> This question and subsequent similar questions you are asked to think about the extreme behaviours of individuals in your school.
<sup>13</sup> It seems likely to us that there will be little difference in the behavior and management regime for very young children but this is an opportunity for you to disagree.
<sup>14</sup> Numbers, character and organisation of year 2 to 5 (emergency workers and vulnerable children) may differ from year 6 affecting contacts. This question might not be relevant to some schools

|  |  |
| --- | --- |
| 8b | *Thinking about the behaviour of individual Cohort 3 adults, give a range of contacts for Cohort 3 adults around your central answer to Q8a for <u>normal</u> times.* |
| 9a | *How many people (both children and other adults) does a Cohort 3 adult come into contact (within 1 metre for 5 minutes or more) with on a <u>new normal</u> school day??* |
| 9b | *Thinking about the behaviour of individual Cohort 3 adults, give a range of contacts for Cohort 3 adults around your central answer to Q9a for <u>new normal</u> times.* |
| 10a | *How many people (both adults and children) does a Cohort 4 adult come into face to face contact (within 1 metre for 5 minutes or more) with on a <u>normal</u> school day?* |
| 10b | *Thinking about the behaviour of individual cohort 4 adults give a range of contacts around your central answer to Q10a for <u>normal</u> times.* |
| 11a | *How many people (both adults and children) does a Cohort 4 adult come into face to face contact (within 1 metre for 5 minutes or more) with on a <u>new normal</u> school day?* |
| 11b | *Thinking about the behaviour of individual cohort 4 adults give a range of contacts around your central answer to Q16 for <u>new normal</u> times.* |
| 12 | *How many children does **an adult (from both Cohort 3 and 4)** have physical/face-to-face contact (within 1 metre for 5 minutes or more) during a typical day, in <u>normal</u> times?* |
| 13 | *How many adults does **an adult (from both Cohort 3 and 4)** have physical/face-to-face contact (within 1 metre for 5 minutes or more) with during a typical day, in <u>normal</u> times?* |
| 14 | *How many children does **an adult** have physical/face-to-face contact (within 1 metre for 5 minutes or more) with during a typical day, in <u>new normal</u> times?* |
| 15 | *How many adults does **an adult** have physical/face-to-face contact (within 1 metre for 5 minutes or more) with during a typical day, in <u>new normal</u> times?* |
| 16 | **Adherence.** *All schools will seek to provide a safe environment following guidelines (e.g. DFE) where feasible and from their own management decisions. Of course some recommendations and procedures may be easier to adhere to than others. We would like your assessment of the extent to which your school is able to follow guidelines. 100% means perfect adherence while 0% means no adherence at all. We have left some space below if you want to make any comments on the any factors that make it difficult to reach 100%* |
| 20 | *Weather might affect the ability to socially distance and being outdoors is recognised as being less risky than indoors. Estimate the increase in number of contacts as a percentage increase among children as a consequence of bad weather. (e.g. an answer of 30% will indicate the number of contacts increase by this percentage due to play breaks being inside)* |
| 21 | *How many times a day do your children wash their hands? Give a range if applicable* |

Table 4 list the results, noting that triplets of quantiles for the elicitation of a single central are variance spreads on the single values, rather than the usual elicited uncertainty ranges per expert. In several questions the experts are asked a pair of questions to estimate contact numbers for an individual (Q2a, Q3a, Q5a,Q6a) and then to requested to estimate the range of contacts considering the variations between individuals (Q2b, Q3b, Q5b, Q6b). Figure 3 shows examples of responses to some questions to illustrate variation in responses among the experts. Figure 4 shows the range graphs for the DM contact distributions, while Figure 5 shows a typical distribution for an individual child. The amalgamated DM distributions are characteristically heavy tailed. Figure 6 plots the 5^th^, mean and 95^th^ values to compare normal (pre-COVID) and new normal (COVID) times.

**Table 4.**
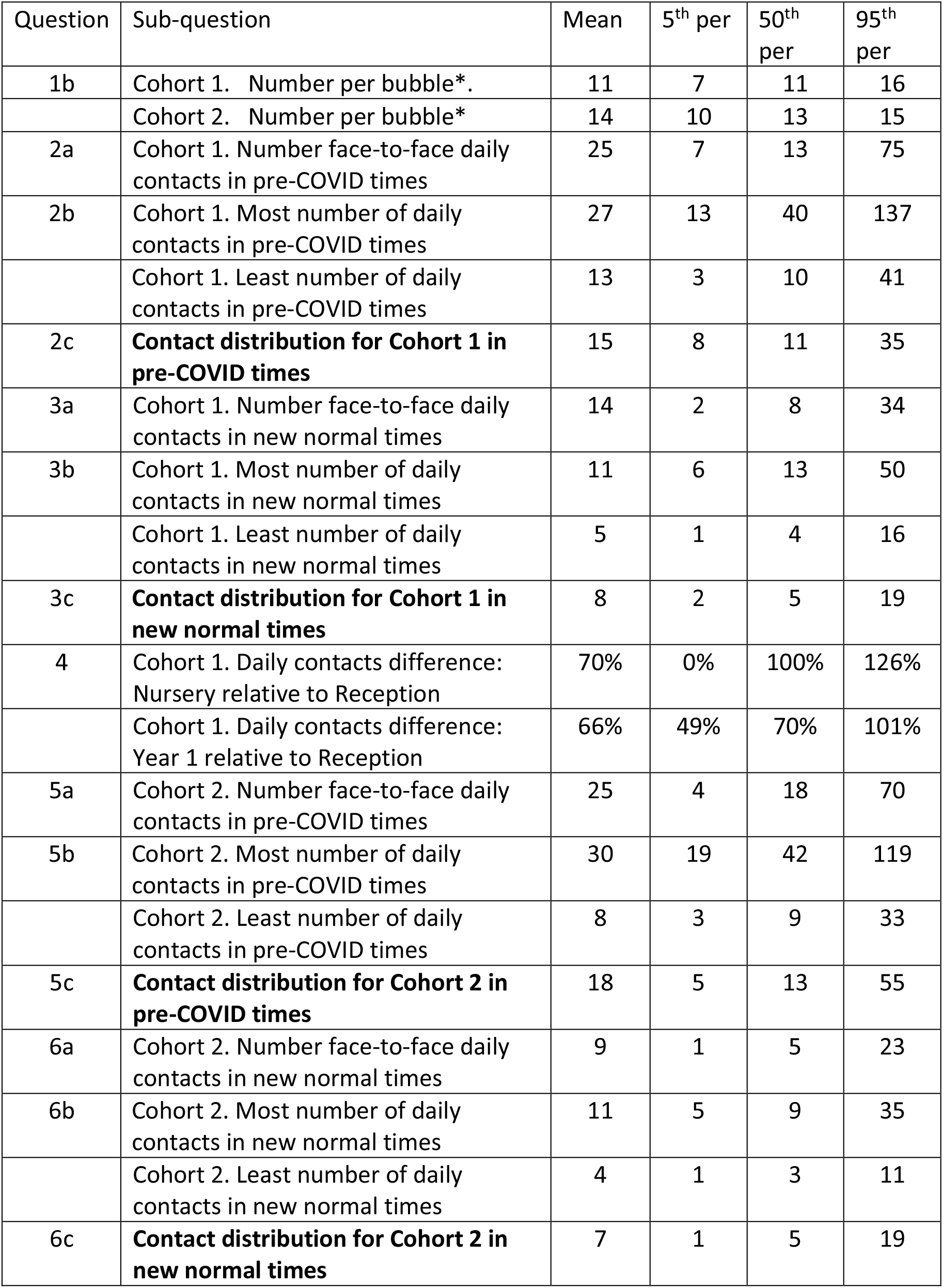
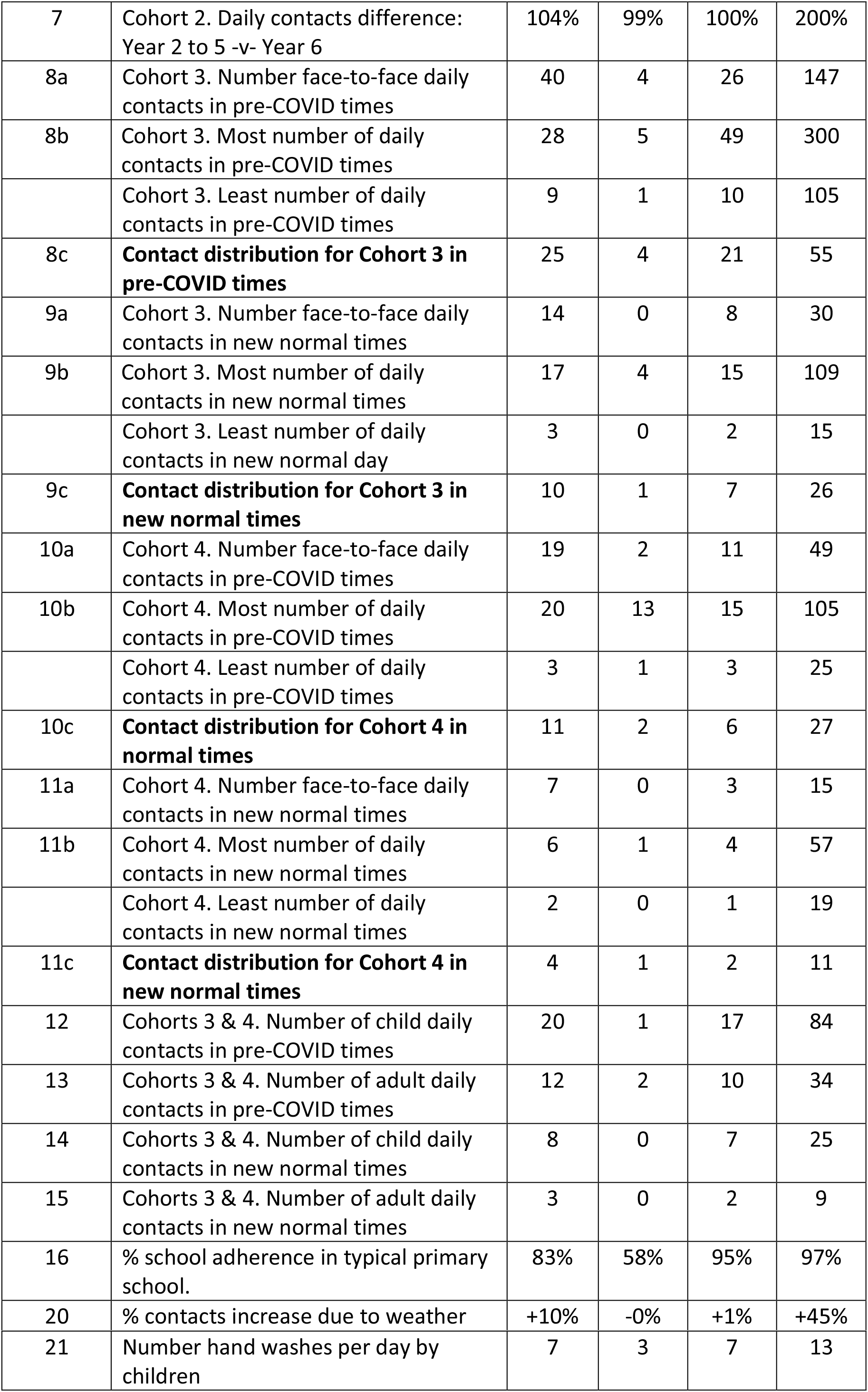
Results of Elicitation. Unless otherwise indicated the data are contacts per day. * denotes represent 5^th^, 50^th^ and 95^th^ percentile variance spreads on the single values, rather than the usual elicited uncertainty ranges per expert. Contact distributions are shown in bold as these are judged to be the most appropriate for epidemiological transmission modelling.

| Question | Sub-question | Mean | 5<sup>th</sup> per | 50<sup>th</sup> per | 95<sup>th</sup> per |
| --- | --- | --- | --- | --- | --- |
| 1b | Cohort 1. Number per bubble*. | 11 | 7 | 11 | 16 |
|  | Cohort 2. Number per bubble* | 14 | 10 | 13 | 15 |
| 2a | Cohort 1. Number face-to-face daily contacts in pre-COVID times | 25 | 7 | 13 | 75 |
| 2b | Cohort 1. Most number of daily contacts in pre-COVID times | 27 | 13 | 40 | 137 |
|  | Cohort 1. Least number of daily contacts in pre-COVID times | 13 | 3 | 10 | 41 |
| 2c | **Contact distribution for Cohort 1 in pre-COVID times** | 15 | 8 | 11 | 35 |
| 3a | Cohort 1. Number face-to-face daily contacts in new normal times | 14 | 2 | 8 | 34 |
| 3b | Cohort 1. Most number of daily contacts in new normal times | 11 | 6 | 13 | 50 |
|  | Cohort 1. Least number of daily contacts in new normal times | 5 | 1 | 4 | 16 |
| 3c | **Contact distribution for Cohort 1 in new normal times** | 8 | 2 | 5 | 19 |
| 4 | Cohort 1. Daily contacts difference: Nursery relative to Reception | 70% | 0% | 100% | 126% |
|  | Cohort 1. Daily contacts difference: Year 1 relative to Reception | 66% | 49% | 70% | 101% |
| 5a | Cohort 2. Number face-to-face daily contacts in pre-COVID times | 25 | 4 | 18 | 70 |
| 5b | Cohort 2. Most number of daily contacts in pre-COVID times | 30 | 19 | 42 | 119 |
|  | Cohort 2. Least number of daily contacts in pre-COVID times | 8 | 3 | 9 | 33 |
| 5c | **Contact distribution for Cohort 2 in pre-COVID times** | 18 | 5 | 13 | 55 |
| 6a | Cohort 2. Number face-to-face daily contacts in new normal times | 9 | 1 | 5 | 23 |
| 6b | Cohort 2. Most number of daily contacts in new normal times | 11 | 5 | 9 | 35 |
|  | Cohort 2. Least number of daily contacts in new normal times | 4 | 1 | 3 | 11 |
| 6c | **Contact distribution for Cohort 2 in new normal times** | 7 | 1 | 5 | 19 |
| 7 | Cohort 2. Daily contacts difference: Year 2 to 5 -v- Year 6 | 104% | 99% | 100% | 200% |
| 8a | Cohort 3. Number face-to-face daily contacts in pre-COVID times | 40 | 4 | 26 | 147 |
| 8b | Cohort 3. Most number of daily contacts in pre-COVID times | 28 | 5 | 49 | 300 |
|  | Cohort 3. Least number of daily contacts in pre-COVID times | 9 | 1 | 10 | 105 |
| 8c | **Contact distribution for Cohort 3 in pre-COVID times** | 25 | 4 | 21 | 55 |
| 9a | Cohort 3. Number face-to-face daily contacts in new normal times | 14 | 0 | 8 | 30 |
| 9b | Cohort 3. Most number of daily contacts in new normal times | 17 | 4 | 15 | 109 |
|  | Cohort 3. Least number of daily contacts in new normal day | 3 | 0 | 2 | 15 |
| 9c | **Contact distribution for Cohort 3 in new normal times** | 10 | 1 | 7 | 26 |
| 10a | Cohort 4. Number face-to-face daily contacts in pre-COVID times | 19 | 2 | 11 | 49 |
| 10b | Cohort 4. Most number of daily contacts in pre-COVID times | 20 | 13 | 15 | 105 |
|  | Cohort 4. Least number of daily contacts in pre-COVID times | 3 | 1 | 3 | 25 |
| 10c | **Contact distribution for Cohort 4 in normal times** | 11 | 2 | 6 | 27 |
| 11a | Cohort 4. Number face-to-face daily contacts in new normal times | 7 | 0 | 3 | 15 |
| 11b | Cohort 4. Most number of daily contacts in new normal times | 6 | 1 | 4 | 57 |
|  | Cohort 4. Least number of daily contacts in new normal times | 2 | 0 | 1 | 19 |
| 11c | **Contact distribution for Cohort 4 in new normal times** | 4 | 1 | 2 | 11 |
| 12 | Cohorts 3 & 4. Number of child daily contacts in pre-COVID times | 20 | 1 | 17 | 84 |
| 13 | Cohorts 3 & 4. Number of adult daily contacts in pre-COVID times | 12 | 2 | 10 | 34 |
| 14 | Cohorts 3 & 4. Number of child daily contacts in new normal times | 8 | 0 | 7 | 25 |
| 15 | Cohorts 3 & 4. Number of adult daily contacts in new normal times | 3 | 0 | 2 | 9 |
| 16 | % school adherence in typical primary school. | 83% | 58% | 95% | 97% |
| 20 | % contacts increase due to weather | +10% | -0% | +1% | +45% |
| 21 | Number hand washes per day by children | 7 | 3 | 7 | 13 |

**Figure 3.**
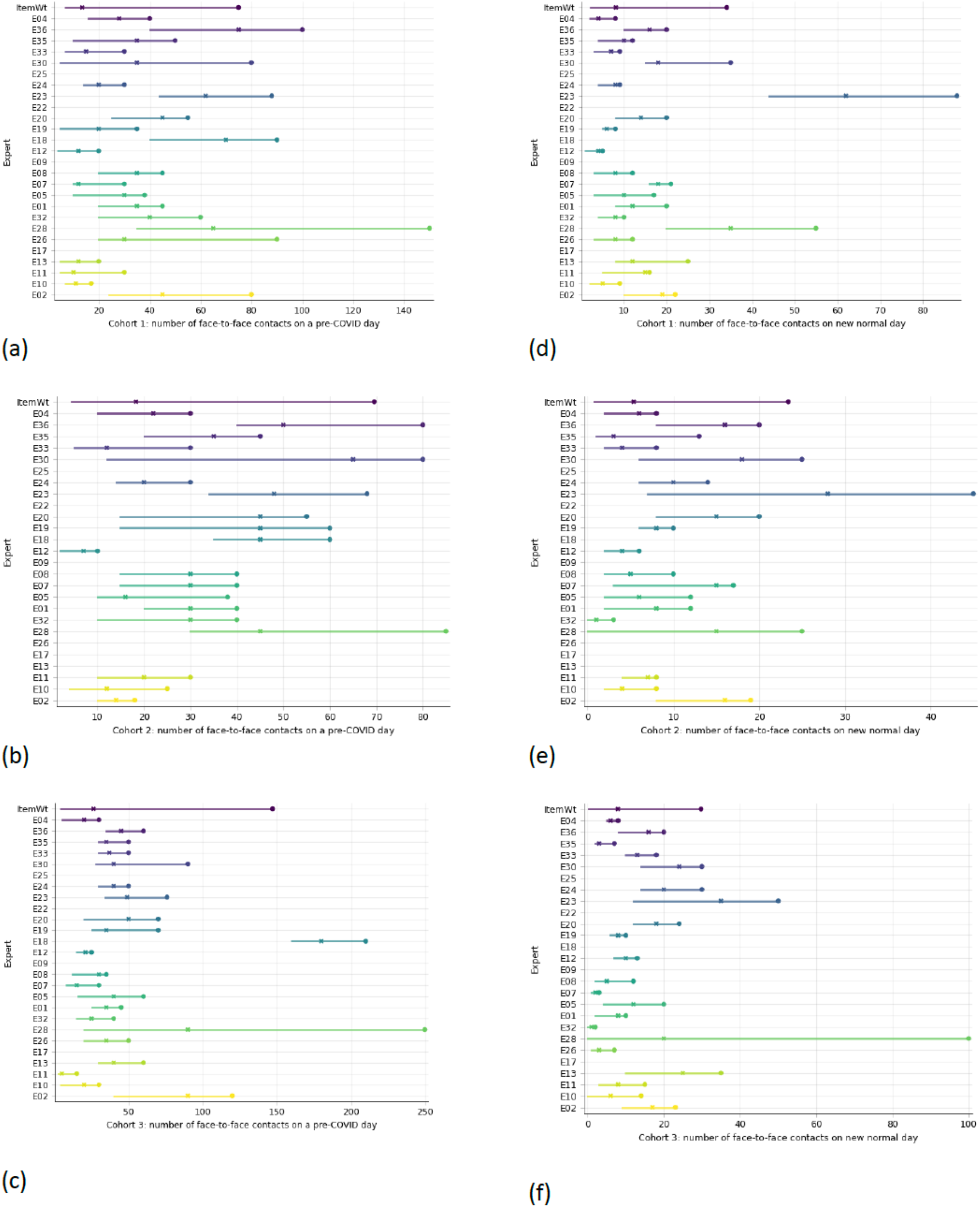
Expert range graphs for (a) Q2a (b) 5a, (c) 8a, (d) 3a, (e) 6a and (f) 9a.

**Figure 4.**
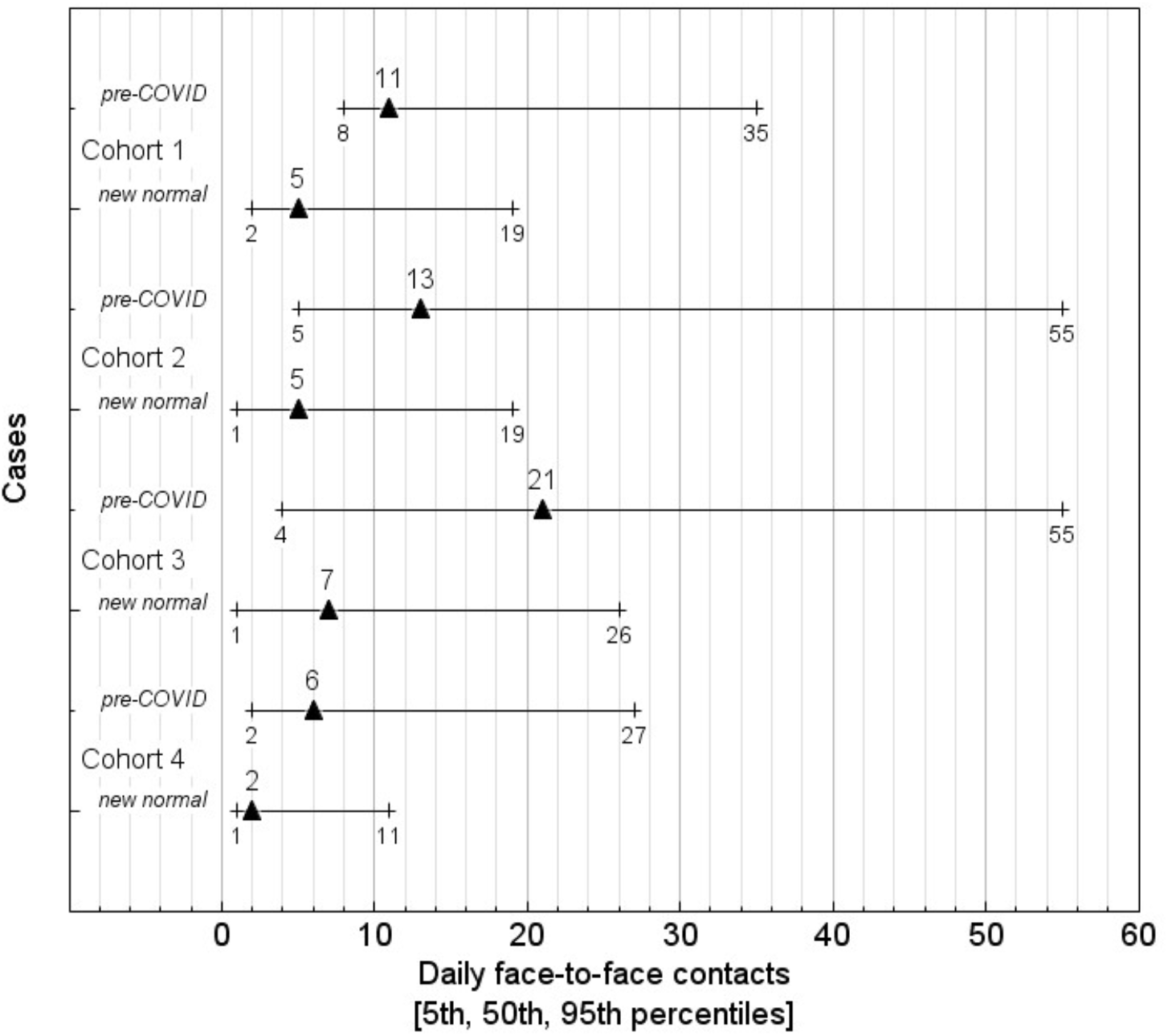
shows the DM range graphs for the daily contacts for the four cohorts.

**Figure 5a.**
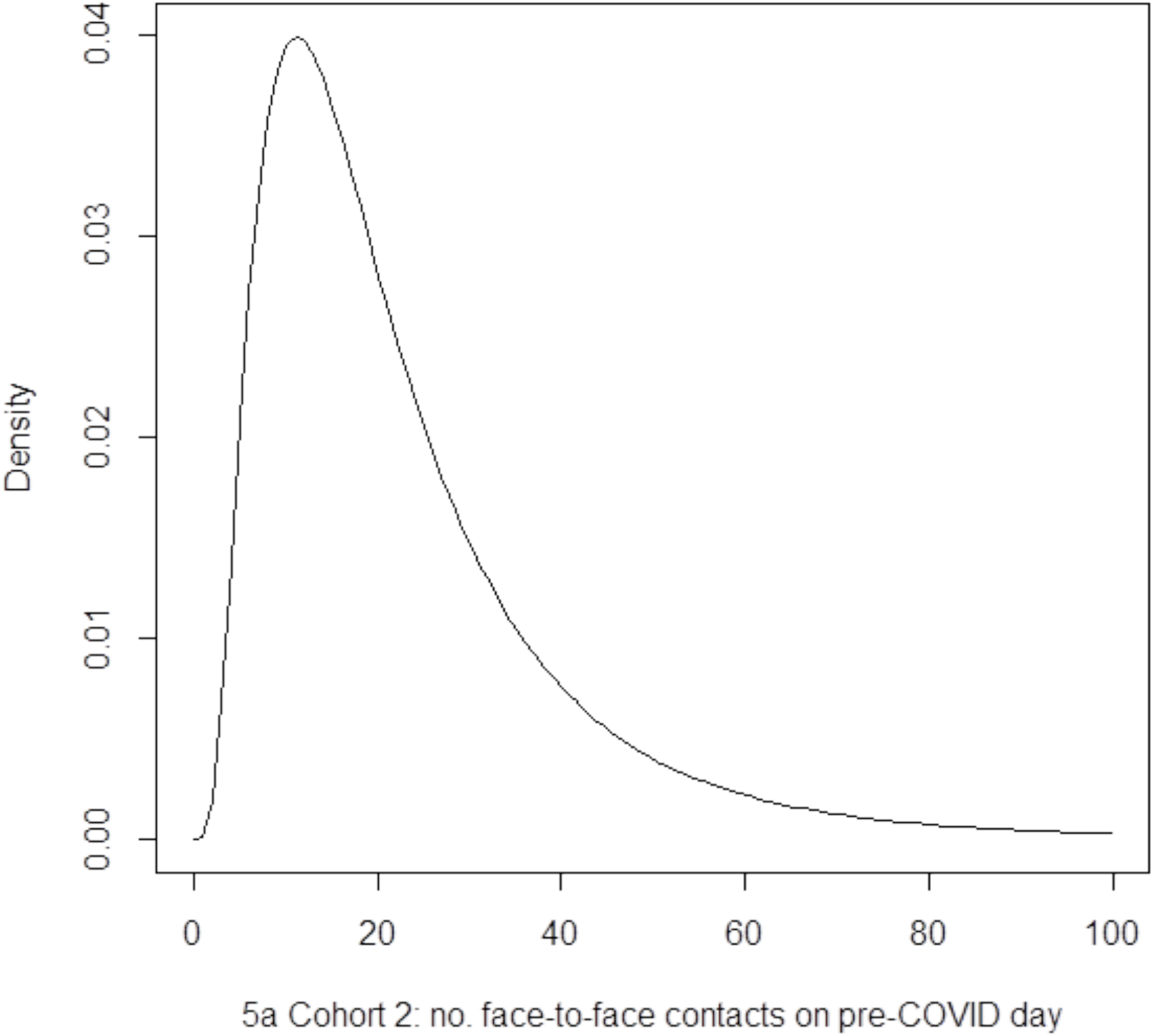
Decision Maker distribution for contacts of a typical individual child from Cohort 2.

**Figure 6.**
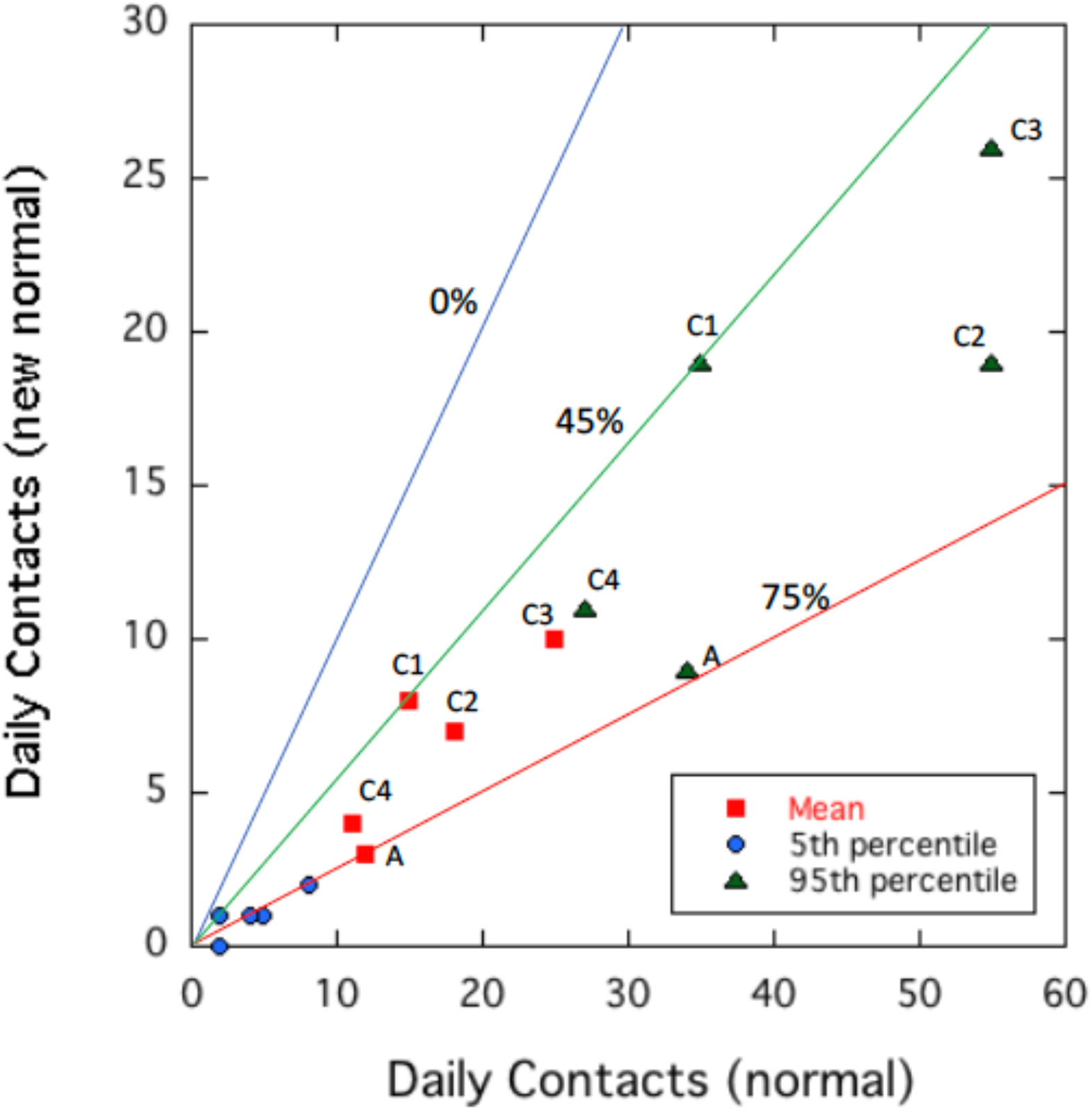
Plot of 5^th^ ile, mean and 95^th^ ile daily contacts for normal (pre-COVID) and new normal (COVID) times. The four cohort elicited data for the mean and 9^th^ ile are labelled. A represents adult to adult contacts in the school. The lines represent no change in contacts while the two other lines are for constant percentage reductions in daily contacts that bound the elicited data.

The interviews conducted suggested the teachers were largely thinking about how contacts might vary from day to day, with this being much harder to estimate during normal times due to the diversity of activities pursued by adults and children. For the second set of questions the variation between experts was also dependent on the make-up of the school; for example some schools had special-learning units which meant some children in the cohort spent only part of the day engaged in mainstream learning.

Before presenting the responses to specific questions we consider sources of uncertainty in the results with a focus on the contact data. Some questions concern the contacts of individuals while others concern variations among groups of individuals. Each teacher has been asked effectively to make measurements of contacts. The definition of a contact (conversation at 1 metre for five minutes or more) is challenging and quite large uncertainty is implicit in making the measurement. The large range for most experts in the graphs for individuals (Figure 3) represents the accuracy of the measurement. In the interviews teachers also placed different emphasis on thinking this through; in some instances 5 minutes of contact was considered a long time for some children during the course of play-led learning.

In comparing schools and combining experts into a DM two additional sources of uncertainty arise. First there are likely to be real differences between schools because of variations in bubble sizes and school characteristics. In all of the interviews the teachers attested to the strict way in which the bubbles were observed, and thus influenced their thinking around contacts. Second there is a calibration issue with each expert working out how to make the measurement. These uncertainties are manifest in the wide variation of elicited values observed in Figure 3. In normal circumstances face-to-face discussions between experts might have been able to reduce the calibration effect. In creating the DM we are combining measurement accuracy at schools, real variations in contacts between schools and significantly different calibrations.

The DM distribution aggregates all these uncertainties and results in markedly skewed distributions (Figure 4a). Here we take the ratio of the right to the left tail, commonly termed eccentricity, as an approximate measure of skewness. Table 5 compares the average eccentricity of the individual experts with the DM eccentricity for several questions. The conflation of all the uncertainties with a DM distribution leads to a quantitative description of the uncertainty of the contacts typical or average pupil, but because it integrates measurement errors, calibration errors and aleatory uncertainty it is not appropriate for developing an epidemiologically relevant model of contacts.

**Table 5.** Ratio of right to left tail (eccentricity) as proxy for skewness contrasting average of individual experts and Decision Maker

| Question | Expert ratio | DM ratio |
| --- | --- | --- |
| 2a | 1.86 | 9.36 |
| 3a | 1.29 | 4.45 |
| 5a | 1.30 | 3.71 |
| 6a | 1.18 | 5.52 |
| 8a | 1,71 | 8.64 |
| 9a | 1.42 |  |

**Table 6.** Questions related to risk mitigation.

| Question |  |
| --- | --- |
| 1a | *Please describe your strategy to reduce close contacts between pupils (about 50 words). You might want to discriminate between Cohort 1 and 2 above.* |
| 18 | *Describe briefly the cleaning regime in your school using disinfectant (maximum 50 words)* |
| 19 | *Describe the policy for parents to drop off and pick up their children each day (50 words maximum).* |
| 20 | *Weather might affect the ability to socially distance and being outdoors is recognised as being less risky than indoors. Estimate the increase in number of contacts as a percentage increase among children as a consequence of bad weather. (e.g. an answer of 30% will indicate the number of contacts increase by this percentage due to play breaks being inside)* |
| 21 | *How many times a day do your children wash their hands? Give a range if applicable* |
| 22 | *What is your policy if a child or adult staff develops possible COVID19 symptoms (either outside school or during school hours) (50 words maximum)?* |
| 23 | *What is your policy if a parent or another relative of a child contracts COVID19?* |
| 24 | *Other Comments. Please make any remarks about additional risk factors that you think should be considered (maximum 100 words).* |

Epidemiological models of transmission convert contact data into a distribution of individual R_0_ values (Brooks-Pollock et al. 2020b). Here we therefore have created a DM distribution from data on the median contacts that characterize the average person (Q2a, Q3a, Q5a, Q8a, Q9a, Q10a and Q11a) combined with least and most number of contacts as tails (Q2b, Q3b, Q5b, Q8b, Q9b, Q10b and Q11b). We interpret least and most as 1%ile and 99%ile values. The interviews confirmed that some teachers worked through the 5 and 95 percentiles by thinking of the most extreme values and then adding even more to account for uncertainty. Thus each expert’s original face-to-face (a) median is coupled with his or her least & most (b) values, with the latter taken as 1%ile and 99%ile values. All experts inputs are processed to form the joint [1; 50; 99] DM for these items. The DM distribution is inspected for its 50 %ile value, and for the corresponding 5 %ile and 95 %ile values, and these are the (c) values reported in Table 4 as Q2c, Q3c, Q5c, Q8c, Q9c, Q10c and Q11c, plus their means.We propose that these distributions are the most appropriate for transforming into distributions of individual R_0_ values for input to epidemiological models.

### Bubbles

An important risk mitigation strategy [Q1b] is to form groups of children (bubbles) who are organized to learn together to limit contacts with many children. The DFE guidelines issued during the study period were for schools to form bubbles of up to 15 children. The responses indicated bubbles between 6 and 15 with a mean of 11 (Cohort 1) and 13 (Cohort 2). The response to the question on spacing between bubbles indicated that each bubble was in a separate room and so these results are not reported as they are deemed not relevant to the risk of transmission between children in different bubbles during class time. Teacher interviews substantiated this view, with teacher-teacher contacts between bubbles being substantially restricted. The interviewees cited this change of behaviour as one of the most significant in terms of reduction in contacts for teachers.

### Contact data

We first consider results for contacts of children in normal pre-COVID times (Q2, Q5). Taking the mean values of 25 contacts for Cohort 1 (Q2a) and 18 contacts for Cohort 2 (Q5a) suggests a minimum of 2.1 and 1.5 contact hours. These values are 30-21% of a typical school day (~7 hours). The distributions are highly asymmetric with high tail extending to high contact numbers. This feature shows up both in the upper end of the distribution for one child (Q2a and Q5a) and for the values for the most contacts (Q2b and Q5b). The combined central values (Q2c and Q5c) reflect variations among children indicating that more active child is 2.3 times (Cohort 1) and 3 times (Cohort 2) more active and the least active child 0.5 (Cohort 1) and 0.3 (Cohort 2) less active than a typical (mean) child. The results for Cohorts 1 (Q2) and Cohort 2 (Q5) are similar with a weak indication that younger children have slightly greater contacts compared to older ones and that older children have a somewhat wider range of activity.

Although our approach is fundamentally different to previous contact survey approaches, we find comparable results. Datasets on primary age children are sparse and often out-of-date. Parents are typically surveyed, although those with children can be included with tailored surveys. The most widely used survey, POLYMOD (Mossong et al. 2008), gives an average of 14.81 (standard deviation of the mean = 10.89) contacts per day for the 5-9 age group (N = 661). The Social Contact Survey, conducted in 2010, collected data within schools from 166 children aged 5-9 years. There, a median of 12 contacts per child was measured (mean 14.6, standard deviation 13.7, 95% percentile range 4, 40) (Danon et al 2013). Other studies (e.g. Zhang et al. 2020) are based on smaller sample numbers (a few tens) for primary age children and more recent UK surveys do not include people under 18 years of age at all (Jarvis et al 2020; Klepac et al 2020). The data in these studies suggest large variations from the mean and a long tail of high contact individuals. These studies do not report 5^th^ and 95^th^ percentiles on the contact distributions. The teacher elicited data for pre-COVID times (Q2c and Q5c) are qualitatively consistent with previous studies.

The contacts between children in new normal time under risk mitigation regimes are substantially reduced (Figure 6): comparing mean contact numbers are reduced by 53% for Cohort 1 (Q3a) and 62% for Cohort 2 (Q6a). The latter reduction is slightly less than the reduction of 74% for adults found by Jarvis et al. (2020). The somewhat greater reduction in contacts in Cohort 2 compared to Cohort 1 in new normal times, however, indicates that older children are slightly easier to manage with respect to social distancing. This interpretation was supported by interviewees. Some thought that older children were more capable of more sustained (> 5m) social distancing (talking face-to-face) in free time, enhanced by reduced contacts engendered by the more formal layout of the school environment. Young children needed more direct contact due to the need for comfort or due to accidents. Long tails indicative of a small number of very active children are still apparent (Q3c and Q6c), but the tails represented by 95^th^ percentile values are reduced by similar amounts to the means. Division into bubbles where class sizes were reduced by factors of 2 and 3 are interpreted to be significant factors in reduction of contacts, corroborated at interview. Much larger decreases in contacts among children of between 4 and 10 are documented in Zhang et al. (2020) as a consequence of lockdown. In this case schools were closed and strict lockdown regimes prevented mixing of children from different families.

The formation of bubbles, however, is unlikely to be the only factor in the decrease in contacts. Children will naturally form smaller groups in schools due to either juxtaposition in class or formation of friendship groups that are much smaller than a classroom (Conlan et al 2011). Thus most contacts will be restricted to a fraction of a class in normal times, while in bubbles the size of friendship groups will become more comparable. The interviews indicated, however, that reception classes in particular were more free flow with gregarious children interacting with sibling groups in other years as well as their peer group within bubbles. These observations help explain the significantly smaller reduction in contacts in Cohort 1.

We asked questions about differences between different cohorts (Q4 and Q7). The responses indicate only weak differences. The data indicate no difference between nursery and reception age children and a weak indication that Year 1 children have slightly fewer contacts. However, the uncertainties are large indicating low confidence in these judgements. Year 6 are not judged to be different to Years 2 to 5.Views about this were controlled by the idea that children’s behaviour changed gradually and that within any one peer group this varied between children. Nonetheless there was a fairly uniform view that there was greater potential to mitigate contact behavior in older children, that changes in class layout had a strong control on ‘in class’ contacts, and that younger children tended to play side by side but need more close contact with adults. <u>The</u> view that younger children are more difficult to manage than older children is supported though by the responses the greater reduction in contacts for Cohort 2.

The responses indicate that Classroom staff (Cohort 3) have a mean total contact number of 25 with both children and adults (Q8c). Typically classes in primary school involve 30 pupils with a single teacher and the same teacher will have *ad hoc* interactions with other teachers and staff during the day. Converted into duration of contacts this equates to a minimum of 2.2 contact hours. Ranges of contacts among teachers (Q8c) indicate a large variation. Again the results indicate some skewness with a heavy tail of large contact numbers. The proportion of children and other adults can be evaluated from Q12 and Q13, noting that the total mean number of contacts from the answers of 32 (Q12 + Q13) can be compared to 25 (Q8c) indicate consistent responses. The results indicate contacts involve 65% children and 35% adults.

Approximating contacts for adults (in the expected age ranges of teachers and teaching assistants) are reported to be about 12-14 per day (Mossong et al. (2008) and approximately 15-20 per day (Zhang et al. 2020). In the Social Contact Survey, 298 participants were teachers: they reported a median of 18 and mean of 51.2 contacts per day (Danon et al 2013). The mean of 25 daily contacts in our study is consistent with the Social Contact Survey results, taking account that school hours are only a part of the day. The high number of contacts within school hours for normal times indicates the interaction with their charges.

In new normal times mean daily contacts for Cohort 3 (Q9c) decrease from 25 to 10, a 60% reduction (Figure 6). The reduction in contacts compared to normal times is a factor of 2.5, compared to between 2.1 (Cohort 1) and 1.5 (Cohort 2) for children. Note that the mean of 10 is similar to the bubble size of 11-13 (Q1b). The contacts between adults between normal (Q13) and new normal (Q15) reduces by 80% (Figure 6). This result is comparable to a 74% reduction in adults found by with Jarvis et al (2020).

These results indicate that teachers are social distancing to a greater extent than children, limiting close contacts as far as is feasible. Answers to Q9b indicate variations in the roles of different teachers and effects of tasks like supervising breaks and meal times. There is still a heavy tail to the responses but this is not as great as for other responses. In the interviews the teachers perceived that those facing older children were strongly socially-distanced but those facing younger children were less able to strictly observe this.

For Cohort 4 the changes between normal (Q10) and new normal times is similar to Cohort 3, but in general contacts are fewer (by ~ 30%) and reflect different roles of ancillary staff some of which involved much less interactions with children (e.g. administrative staff). Responses to Q11 indicate a 64% reduction of contacts in new normal time (Figure 6), reflecting the deliberate policy of limiting ancillary staff contacts with children and the efforts of these staff to observe social distancing. Adult to adult contacts are reduced by 80% between normal and new normal time (Figure 6). At interview Cohort 4 were uniformly perceived as adults with the greatest change to their day-to-day contacts in the working day, with some able to be entirely socially distant. This is similar to the overall reduction in contacts of 20% in the general community reported in Brooks-Pollock (2020a). Inevitably a school is a work place where overall adult to adult contacts are inevitable; the daily contacts within school hours are at the lower end of contact numbers reported in other contact studies (Mossong et al. 2008; Danon et al. 2013; Zhang et al. 2020).

In Figures 4 and 6 the tails of the contact numbers (5^th^ and 95^th^ percentiles) are compared and the overall reductions in contacts are similar to the mean falling within the 45%-80% range. The comparison reduction in contacts due to the risk mitigation arrangements are approximately as effective across the wide range of daily contacts of individual children, classroom staff and support staff.

We asked about adherence and the response indicates that the experts considered adherence to be high (Q16). This view is supported by the analysis of the qualitative parts of the questionnaire related to risk mitigation measure. From the interviews the string was driven often by thinking about consequences, for children, their families but also their colleagues. Questions related to understanding risk reduction (Q20, Q21, Q22) are discussed in the next section.

## Risk Mitigation Survey Results

We asked the Head Teachers about risk mitigation measures that they have put in place. Questions are listed in Table 4. Reference is made for some topics to literature on the quantification of the risk being mitigated. Most questions answered, from 23 teachers, except Q1a (21) and Q24 (15). Verbatim individual responses are provided in appendix 2 where many interesting points and examples of innovation in risk management, mitigation and reduction are apparent. To augment the elicitation and survey question answers structured interviews were conducted to help in the interpretation of the response.

Teachers have attempted to follow the Government guidelines, with respect to school reopening. Links to guidelines from DfE can be found as footnotes. These guidelines have been updated regularly so it is not easy to categorically state which guidelines were followed when teachers completed their elicitations. It is apparent that in instances where the guidelines are not defined, such as ‘minimising contact and mixing by altering, as much as possible…’ teachers have typically been very cautious, putting in measures that go beyond the guidelines. Equally, there are no explicit guidelines related to the need to ‘wash hands frequently’ and consequently there is considerable variation in interpretation. One interviewed teacher made the point that the younger children are in the process of learning, and that learning needs to incorporate the making of mistakes. Many schools had made a good job of initiating procedures for handwashing that were age appropriate and encouraged sticking within the guidelines (e.g. no singing). Teachers have interpreted the guidelines to suit their own settings and then gone one step further to ensure the safety of their charges. Further comparisons to the guidelines are included in each section below. The appendix reports all responses and provides a record of good practice and innovation by the schools which will shared between the schools.

Answers to Q1a concerning the strategy to reduce close contacts of children were broadly similar for cohort 1 and 2 and have been combined for this summary. Four responders did not answer this question. The Government <u>guidelines</u> recommend smaller group sizes, with consistent students and teachers, kept 2 m apart where possible with staggered break times. Outdoor activities are recommended where possible and students are to stay in the same desks and rooms as much as possible. 76% of responders noted social distancing measures that were being put in place with visual indicators for children to follow. Over 90% of responders followed the guidelines and indicated the use of classroom bubbles of varying sizes below the recommended number of 15. The answer to Q1b (Table 7) indicated that many bubbles were about 10 children. Some have indicated that children will only be in school part time to allow teachers to accommodate the reduced class sizes.

Other measures included the removal of furniture to allow for more space, rotation of toys and/or removal of certain play items and lunches taking place in the classroom (either packed lunches or ‘take-away’ style cartons to remove the need for cutlery). In line with the guidelines, a third of the responses indicated that learning would be moved outdoors as much as possible and a similar number noted the need for individual desks and resources. Around a third of teachers have extrapolated the guidelines and ensured that student bubbles are allocated their own areas of the playground, their own toilets or their own lunchtime spaces and over 50% refer to the suggested staggered break times, start times etc. This was also borne out at interview, almost all interviewees used the word ‘strict’ to describe their bubble, and indicated that adults avoided connecting between bubbles.

Unsurprisingly all responders in Q18 concerning cleaning have referred to changes to the usual cleaning regimes in their school with over 70% making specific reference to ongoing cleaning throughout the day, highlighting high touch areas such as computers (3 responses) and toilets (13 responses); this is broadly in line with Government recommendations^2^. In the main, deep cleaning is taking place before and/or after school and reference is made to deep cleaning taking place after classroom bubbles have used specific areas; Government guidelines recommend thorough cleaning at the end of the day^3^; 3 responders made specific reference to hard plastic toys being sterilised in Milton after use following DfE guidelines that recommend that cleaning of toys takes place between use^4^. Some teachers referred to the removal of soft toys and furnishings in their answer to question 1a). Three responders have referred to their employment of additional cleaning staff and three have indicated that staff and children will, themselves, engage in cleaning processes. Several responders noted that classroom doors will be left open, presumably to avoid the need to touch door handles as recommended in the Government guidelines^5^. Some of the responses in the appendix indicate that some schools have gone beyond the guidelines, such as: bubbles having their own toilet cubicle and sink; cleaning staff using PPE which are double bagged and stored for 72 hours before putting in bins; disinfecting laptops after every use. Allergies to disinfectant in some of children hampered cleaning of some areas.

SARS-CoV-2 can persist on surfaces for hours to days (Eslami and Jalili 2020) and disinfecting surfaces is recommended by major health authorities (WHO 2020). Exposure via contaminated environmental surfaces appears to be a secondary vector of transmission; transmission via airborne droplets is more important. There is very little direct research on SARS-CoV-2 =so studies rely on tests performed on similar viruses, or other viruses that are hard to kill. Surface disinfection to remove viruses include Quaternary ammonium, ethyl alcohol, hydrogen peroxide and sodium hypochlorite (Henwood 2020). Use of Disinfectants are most effective when used in combination with ultraviolet and vaporised hydrogen peroxide to neutralise aerosolised virus in the air column. Quantifying risk reduction as a consequence of regular deep cleaning is difficult and is notably absent from studies on the role of cleaning (Eslami and Jalili, 2020). If after cleaning virus builds upon surfaces linearly with time to some steady state level after a few hours then cleaning will might reduce the chances of infection through touching contaminated surfaces.

Answers to Q19 were consistent and included implementing staggered times, one-way systems, different entrances, no adults on site and no playground meeting. Other than different entrances for individual groups, these measures follow Government recommendations^6^: 65% of responders indicated a staggered drop off and collection time for children by either year group or class bubbles. 2 further teachers noted the challenges this puts in place for family groups and had amended their policy accordingly. 39% had put in place one-way systems to reduce contact between parent/children arriving and those leaving (at drop off and collection times). 52% noted that parents were either not allowed on the school site or that only one parent could drop the child off, with teachers meeting children at the school gates. 48% referred to social distancing measures being put in place for parents waiting and/or for children in the playground. Approximately 20% referred to children not being allowed in the playground at the start of the school day and needing to go straight to class and a further 20% referred to classes and bubbles having separate entrances and exits.

Accurate quantification of the risk reduction gain from such measures is difficult. A scoping calculation, however, can give an indicative estimate. If the average parent has a daily contact hours of 30 (Danon et al. 2013) then the mitigation measures described might reduce the contact hours by 1 or 2 hours compared to normal mixing associated with delivering children to and from school, so we estimate an overall effect of a few percent (~3-6%), which can be compared to the 80% reduction associated with general lockdown (Brooks Pollock et al. 2020). The contribution to risk reduction within schools is likely to be very small, but larger and tangible in the wider community.

Responses to the question on weather (Q20) indicate that contacts were not changed between indoors and outdoors. It is now widely thought that outdoors is much less risky than indoors but this was not the question. Interviews indicated that the physical layout of the school informed their responses, and some commented the influence of very good weather during the course of the study.

In relation to handwashing (Q21) Government guidelines recommend that adults and children should frequently wash their hands with soap and water for 20 seconds and dry thoroughly^7^. Responses (Table 7) gave a range from 3 to 13, with over half opting for a range between 3 and 10 handwashes. One teacher referred to the number of handwashes being dependent on the number of visits to the toilet. Those teachers who gave a specific lower number with a wider range have implemented a policy of handwashing at certain times with variance added for toilet visits (one responder has indicated scheduled handwash times). All responses are shown in the elicitation results.

There is limited evidence on the efficacy of handwashing in reducing transmission. Soap is very effective at neutralising the virus (Eslami and Jalili, 2020). The chances of an infectious individual passing the virus by touching a person or contaminating a surface are reduced by regular hand washing. The weight of evidence is that handwashing reduces transmission but may become counterproductive, less effective and even harmful if the handwashing becomes excessive. Risk reduction is estimated as 6% to 44% for respiratory diseases (Rabie and Curtis, 2006). Beale et al (2020) studied risk reduction of handwashing for flu and common coronavirus infections in the UK. They estimate significant reduction for between 6 and 10 handwashes per day (5^th^ per = 58%; 50^th^ per = 36%; 95^th^ per = 1%), results consistent with those of Rabie and Curtis, 2006). A reservation for these studies is that they involve extrapolation to transmission of SARS-CoV-2. This distribution is included in the risk modelling. The range in the elicited answers indicate that most schools have adopted an optimal regime of hand washing too reduce risk and have avoided excessive handwashing that are considered to be potentially harmful.

The schools follow Government guidelines for those displaying COVID19 symptoms (Q22), but some schools have gone beyond these recommendations. In general anyone showing symptoms is self-isolated with some mentioning designated areas set aside for this purpose. A few will use of PPE, which is recommended if the adult cannot keep 2 m apart. The symptomatic persons are then sent home immediately and asked to get tested. School areas are then cleaned following the guidelines^8^ to use PPE. Two responders plan to notify families of children in a bubble. One school proposed that everyone in that bubble would be recommended to get a test and, if it comes back positive, the whole bubble is required to self-isolate for 14 days. The individual stays at home for 7 days or until a negative test is achieved. One responder has said that if a positive test is confirmed the whole school will close. For case where infection is suspected outside school hours, those that answered required the school to be informed, a test will be undertaken, and the school notified of the result. A small number mentioned that the person showing symptoms would not be able to attend until a negative test was achieved. One responder stated that a positive test would result in self-isolation of the whole bubble for 14 days. Only a few responded to the policy for dealing with anyone with symptoms identified outside of school. Most interviewees had experienced at least one instance of a suspected case that required testing. While the protocols all required that the child stayed away from school, the responses varied in terms of the notification or isolation of the child’s bubble until test result was received. The DfE guidance states that the bubbles only close if a positive test is returned but one school will require immediate self-isolation until results come back.

The survey indicates that all schools had strong measures in place following and going beyond government guidelines to minimise risk.

Most responders to Q23 stating that the child cannot attend school and must self-isolate - Government guidelines are limited to just this piece of advice, with respect to schools^9^. A small number will inform other families within the same bubble. One school stated they had no policy for such events and 2 schools indicated that the child of said adult could still come to school. One school submitted this guidance from a local health protection team.

Other comments (Q24) are unique to individual schools and are all included in the appendix below. Some noted issues of EAL and SEND students^10^ and risk factors involved with school transport.

The structured interviews augmented these findings from the risk mitigation survey, but highlighted some additional issues. Interviewees mention the difficulties and stress for staff in maintaining social distancing and risk mitigation measures. The tension between social distancing and key educational objectives of learning and developing social skills was highlighted. There was comment that some measures would be impossible or much more difficult with a full return of school.

## Discussion

The circumstances in English primary schools in June and July 2020 are unprecedented and unlikely to be repeated. The partial re-opening of schools was undertaken under strict guidelines of social distancing and a range of risk mitigation methods to reduce the transmission of COVID-19. These unique circumstances provide an opportunity to evaluate the efficacy of different risk reduction strategies and to add to the data on contact patterns for young children and staff in the school environment.

A striking aspect of our results is that the 34 participating schools proved to be representative of schools nationwide. Thus as in opinion polls an accurate picture of what is happening in nationally in schools can be gleaned from a modest sample size. We were fortunate here that volunteerism led to a group of schools covering a wide range of sizes and communities. Even though the schools individually predicted and then reported a very wide range of returns for pupil and teacher attendance on 1^st^ June the average for the 36 schools was very close to the national picture. The prescience of experienced school leaders could be used to anticipate what will happen in September when a full return to school has been mandated by Government.

Breaking social networks is a key non-pharmaceutical intervention for countering the spread of infectious disease. Our study indicates that contacts within schools were reduced in the range 45% to 80% (Figure 5). These strikingly successful outcomes highlight the tremendous work of school staff and the role of greatly reduced class sizes through creation of bubbles of children which are much less than normal class sizes. Although the marked reduction in contacts can be partly explained by much smaller class sizes observations indicate that social groups are characteristically smaller than typical class sizes under normal times (Conlan et al. 2011) Thus organisation of children within bubbles of groups combined with control of their behaviour by teaching staff is a significant factor. A combination of 3/7 of children being invited back to school and limited parental compliance returning eligible children to school on 1^st^ June (30-40%) led to typically only 15-20% of children being present. This allowed schools to form bubble sizes significantly less the 15 DfE guidelines (range 7 to 16 with median of 11). Contact numbers do not scale linearly with group size and the number of contacts saturates at about 20 (Brooks Pollock et al 2020b). The bubble sizes of about 10 have proved to be efficacious in decreasing contacts, limiting mixing between children and thus reducing risk of transmission.

Contacts between adults and children have been reduced by a factor more than 5x between normal and new normal times. The contacts of teaching staff (Cohort 3) decreases by 50% between normal and new normal times indicating that adults can more effectively adopt stringent social distancing practices. A reduction in contacts of about 80% is also approximately the average achieved in the UK at the height of the lockdown (Brooks-Pollock et al 2020b). Given that mixing with groups of children is part of the job of class-room staff the reduction is impressive. Likewise other adult staff achieved similar reductions in contacts. The data also confirm that older children (Cohort 2) are easier to manage than younger children (Cohort 1) with more than a two-fold greater reduction of contacts in the older children compared to the younger ones.

The elicited data can be compared quantitively with other kinds of contact survey data because of the variation in methodologies. The results indicate the same kind of heterogeneity as documented in Danon et al. (2013) in which the data are a mixture of individual contacts involving significant conversations at a distance of 3 m or less or touching plus contacts related to groups where each person in a group counts as a contact. Results were expressed as contacts per day and total contact hours, noting that the latter parameter can exceed 24 hours because contacts within groups occur simultaneously. In a school setting, classroom adults (Cohort 3) have a median of 26 contacts (Q8a), of which about 2/3’s are with children (Q12) and 1/3 with adults (Q13). The distance specified in the question is 1 m with social distancing at the time of elicitation being 2 m. Contact hours are of the order of 2 hours.

In the context of a typical class of 30 the group definition of contacts in Danon et al. (2013) seems less relevant. There must be an additional risk factor related to being in an enclosed space with widespread aerosol circulation, but this is not quantifiable with the elicitation data. However, reducing class sizes from 30 to roughly 10 reduces the random risk of an infectious person being in the class-room by about 1/3.

All of the risk mitigation strategies contribute to reduction of risk. Specific risk management strategies reflect individual circumstances in schools. Many relate to steps which are likely to reduce contacts and disrupt break contact networks. Each of the risk mitigation measures will contribute to risk reduction, but this is hard to quantify.

It seems unlikely that the significant reduction of risk, implied by these results, can be maintained with a full return to school without greatly expanding the accommodation to maintain reduced class sizes, as suggested by the factor of 2 to 3 reduction in contacts between children. Adult staff can continue to observe strict social distancing behaviours and can continue to organise the classroom and break times which reduces contacts.

## Data Availability

Data are in supplementary material material

## Acknowledgements

We are particularly appreciative of the Primary School leaders who volunteered to join the expert panel. Their enthusiasm, support and expert knowledge was paramount. The University of East Anglia fact finding team of Jade Eyles, James Christie, Nicola Taylor and Martin Mangler are much appreciated. The project was completed under the auspices of the RAMP initiative of the Royal Society. Mike Cates is thanked for encouragement and advice.

## Footnotes

1 Optimized weights unweight the lower scoring experts until the performance of the DM in terms of statistical accuracy and informativeness, is optimized

2 https://www.gov.uk/government/publications/coronavirus-covid-19-implementing-protective-measures-in-education-and-childcare-settings/coronavirus-covid-19-implementing-protective-measures-in-education-and-childcare-settings#effective-infection-protection-and-control;

3 https://www.gov.uk/government/publications/coronavirus-covid-19-implementing-protective-measures-in-education-and-childcare-settings/coronavirus-covid-19-implementing-protective-measures-in-education-and-childcare-settings#when-open;

4 https://www.gov.uk/government/publications/preparing-for-the-wider-opening-of-early-years-andchildcare-settings-from-1-june/planning-guide-for-early-years-and-childcare-settings#Section2;

5 https://www.gov.uk/government/publications/coronavirus-covid-19-implementing-protective-measures-in-education-and-childcare-settings/coronavirus-covid-19-implementing-protective-measures-in-education-and-childcare-settings#when-open;

6 https://www.gov.uk/government/publications/coronavirus-covid-19-implementing-protective-measures-in-education-and-childcare-settings/coronavirus-covid-19-implementing-protective-measures-in-education-and-childcare-settings#how-to-implement-protective-measures-in-an-education-setting-before-wider-opening-from-1-june;

7 https://www.gov.uk/government/publications/coronavirus-covid-19-implementing-protective-measures-in-education-and-childcare-settings/coronavirus-covid-19-implementing-protective-measures-in-education-and-childcare-settings;

8 https://www.gov.uk/government/publications/covid-19-decontamination-in-non-healthcare-settings/covid-19-decontamination-in-non-healthcare-settings

9 https://www.gov.uk/government/publications/covid-19-stay-at-home-guidance/guidance-for-households-with-grandparents-parents-and-children-living-together-where-someone-is-at-increased-risk-or-has-symptoms-of-coronavirus-cov

10 EAL are student with English as a second language and SEND are those with special educational needs and disabilities.

11 In this and subsequent similar questions you are being asked about the typical or average behaviour and your uncertainty in this average or typical behaviour.

12 This question and subsequent similar questions you are asked to think about the extreme behaviours of individuals in your school.

13 It seems likely to us that there will be little difference in the behavior and management regime for very young children but this is an opportunity for you to disagree.

14 Numbers, character and organisation of year 2 to 5 (emergency workers and vulnerable children) may differ from year 6 affecting contacts. This question might not be relevant to some schools

